# Low Surgical Skill Rating of Surgeons Increased Complication Rates of Patients after Coronary Artery Bypass Graft

**DOI:** 10.1101/2023.06.19.23291630

**Authors:** Xin Yuan, Kai Chen, Qing Chu, Hansong Sun, Yunhu Song, Sheng Liu, Wei Feng, Xiangqiang Wang, Shuiyun Wang, Liqing Wang, Xin Wang, Fei Xu, Yang Wang, Yanyan Zhao, Shengshou Hu

**Author notes:** These authors contributed equally to this work as the first authors. Address for Correspondence: Shengshou Hu, MD Fuwai Hospital, Peking Union Medical College 167 Beilishi Road, Xicheng District, Beijing, P.R. China.

## Abstract

**Objectives:** It is not well characterized how surgeon-level factors affect surgical outcomes. This study aimed to determine the relationship between surgical skill rating of a cardiac surgeon and complication rates after coronary artery bypass grafting (CABG).

**Methods:** Video-based evaluations of CABG performed by 46 senior surgeons were conducted by 13 blinded expert cardiac surgeons. The evaluations were carried out using a rating score sheet including 7 domains of skills in surgical skills. Other surgeon-level factors such as the demographic information, Myers-Briggs Type Indicator (MBTI) test and the ratio of low frequency and high frequency of heart rate variability (LF/HF) during operation were also collected. The association between surgical skill rating along with surgeon-level factors and surgical outcomes was examined using prospectively collected data. The primary outcome was major adverse cardiac and cerebrovascular event (MACCE), included mortality, myocardial infarction and stroke within 30-day after surgery. The secondary outcome included skill-related complications and all complications. Multivariable generalized linear mixed models were employed for surgical skill rating on MACCE, skill-related complications and all complications.

**Results:** From 2018 to 2021, the participants performed a total of 9,844 isolated CABGs. All participant surgeons were male with a median of CABG volume of 702. Based on three reviewers per video and with a maximum surgical skill score of 5, the average surgical skill scores ranged from 3.10 to 4.81. After adjusted with the risk factors of the patients, compared to the patients of the top quartile rating surgeons, the patients of bottom quartile rating surgeons had 1.381 times of risk in MACCE (95%CI, 1.098-1.735, p=0.006), 1.528 times of risk in skill-related complication (95%CI, 1.340-1.743, p<0.001) and 1.286 times of risk in all complication (95%CI, 1.131-1.463, p<0.001). Surgeons in the top quartile of skill rating had an older age (57.9±4.4 vs. 49.7±5.7, p=0.002), higher rate of “sensing” characteristics of MBTI (72.7% vs. 33.3, p=0.046), lower rate of “thinking” characteristics of MBTI (54.5% vs. 83.3%, p=0.013), higher mean annual volumes of CABG (109.3±57.7 vs. 41.5±25.0 p=0.001), and lower LF/HF (2.3±0.1 vs. 4.9±1.4 p<0.01) than in the bottom quartile of skill rating.

**Conclusion:** Lower surgical skill rating of cardiac surgeon appeared to be associated with higher complication rates for CABGs. The surgical skill rating correlated well with a series of surgeon-level factors. Efforts to improve surgeon surgical skills may result in better patient outcomes and surgeon characteristics may serve as the target of intervention for quality improvements.

## BACKGROUND

Despite the substantial improvements in patient care, cardiac surgery remains a procedure that is highly risky and requires advanced techniques.^1^ Selecting and training competent cardiac surgeons is a challenging and time-consuming, especially with the decreased trainee exposure to cardiac surgery, restricted training hours for resident surgeons, and a shortage of cardiac surgeons. This situation has provoked an increased scrutiny of surgical performance and a need to improve the training process and competence acquisition.^2–6^ Therefore, it is necessary to establish an objective and practical evaluation system to differentiate cardiac surgeons at the same professional level with varying degrees of technical competency.

Surgeon-level factors are important for surgical outcomes but are less studied when compared with factors at patient and hospital levels. Previous studies have suggested that surgeons’ characteristics, such as age, sex, seniority, and volume, are associated with postoperative outcomes.^7–9^ However, the findings in cardiac surgery are inconsistent. Understanding how surgeons’ characteristics impact outcomes is crucial for improving cardiac surgery outcomes. To achieve this purpose, it is crucial to establish a reliable surgical skill evaluation methodology validated using outcomes derived from clinical practice as benchmarks.

Nonetheless, existing studies are lacking with small sample sizes of surgeons and patients, and with a focus on medical students or surgeons in their training stages. Additionally, concurrent surgical skill evaluation tools are generally validated using interrater reliabilities, interstation reliabilities, or surgical faculty rankings.^10–15^

Assuming that variance exists among surgeons who have passed the initial segment of the learning curve, we used data from a single center to determine the surgeon-level factors impacting outcomes for isolated CABG performed by 46 senior surgeons with a total volume of at least 200 cases.

## METHODS

### Participating Surgeons and Surgical Skill Rating

The participating cardiac surgeons were recruited from Fuwai Hospital, a specialized specializing cardiovascular hospital in Beijing, China with an annual surgical volume of approximately 15,000. The study was conducted from January 1, 2018 to April 1, 2022, involving 46 cardiac surgeons.

Each participating surgeon agreed that a researcher would randomly take a video of an isolated coronary artery bypass grafting procedure using the camera in the operating room. The videos were sent to other senior cardiac surgeons from different cardiac centers for surgical skill rating. Thirteen senior cardiac surgeons were invited as raters for the surgical skill rating. Each video was randomly scored by three senior cardiac surgeons. The surgical skill ratings were double-blinded between the participating surgeons and the raters.

A 7-domain rating sheet was constructed to assess surgical skills using a 5-point scale (Table S1).^16^ A score of 1 indicated that the surgical skill was poorly performed during the procedure, while a score of 5 indicated the skill was performed excellently. A score of 3 reflected an acceptable surgical skill of the practicing surgeon. Surgeon raters judged the videos and applied these rating criteria independently. The rating sheet consisted of seven domains in the following areas: (1) Gentleness of movements, (2) Cleanliness of surgical field, (3) Without unnecessary motion, (4) Instrument handling, (5) Coordinate with nurse, (6) Coordinate with first assistant (7) Fluency and coherence of the procedure, as well as an overall skill rating.

The Myers-Briggs Type Indicator (MBTI) test was performed on all participating surgeons via paper questionnaire at the beginning of the study. The MBTI is an inventory that evaluates personality based on the theory of psychological types developed by Carl Gustav Jung. The MBTI classified the personalilty into four demensions represented by a single capital letter (Detailed information in suplementary material).

Heart rate variability (HRV) was test on the participating surgeons during CABG procedure using one 3-lead electrocardiograph (ECG) recording. Three isolated CABG procedures of each participating surgeons were randomly selected for the test. The recording started before surgical hand disinfection and ended after taking off the surgical gown. The exact time of the beginning and end of the anastomotic operation of proximal and distal stomas of the bypass grafts was recorded. The HRV parameters of low frequency (LF) and high frequency (HF) HRV were obtained, and the ration of LF/HF was calculated.^17^

The study was approved by the institutional ethics committee of Fuwai Hospital (IRB2012-BG-006), and written informed consent was provided by participating surgeons. The participants were not financially compensated.

### Surgical Procedures

The isolated coronary artery bypass grafting procedures were recorded on patients aged 18 years or older, performed by the 46 participating surgeons from January 1, 2018 to September 30, 2021. A total of 9,844 isolated coronary artery bypass grafting procedures were included in the study. Detailed clinical data, including baseline characteristics and in-hospital outcomes, were directly collected from the physician order entry and electronic medical record system of Fuwai hospital. The follow-up data was collected by the research staff of Fuwai hospital through telephone interviews.

### Outcomes

The primary outcome was major adverse cardiac and cerebrovascular event (MACCE) included mortality, myocardial infarction and stroke during hospitalization or within 30-days. The secondary outcome included skill-related complications and all complications (the occurrence of any postoperative complication during hospitalization or within 30-days). Surgical complications included mortality, myocardial infarction, hepatic failure, acute kidney injury, stroke, red blood cell infusion, pericardial tamponade, secondary thoracotomy for bleeding, cardiac arrest, ventricular fibrillation, fatal ventricular arrhythmia, III° atrial-ventricular block, intra-aortic balloon pump (IABP) implantation, extracorporeal membrane oxygenation (ECMO) implantation, aortic dissection, coma more than 24 hours, dialysis, re-intubation, re-entry of intensive care unit (ICU), debridement, mechanical ventilation more than 48hours, sepsis, hydrothorax or hydropericardium, pulmonary infection, pulmonary embolism, and atrial fibrillation. Skill-related complications were defined as a composition of post-operative red blood cell infusion, pericardial tamponade, secondary thoracotomy for bleeding, cardiac arrest or ventricular fibrillation, fatal ventricular arrhythmia, myocardial infarction, III° atrial-ventricular block, IABP implantation, and aortic dissection.

The follow-up outcomes included all-cause mortality, myocardial infarction, stroke, chronic kidney insufficiency, re-vascularization, thrombolysis.

### Statistical Analysis

Continuous variables were presented as mean±standard deviation, and categorical variables were presented as frequencies and percentages. The characteristics of the participating surgeons, MBTI test, surgical skill ratings and HRV were determined at the surgeon level, while the outcomes were assessed at the patient level. Analyses accounted for clustering of outcomes from multiple patients treated by a single surgeon. For HRV analysis, the mean value and standard deviation of HF/LF of each participating surgeons were calculated and compared between groups.

For the mean rating of the surgical skill for the participating surgeons, we used the average rating of the seven domains of surgical skill. The seven domains were found to be correlated with each other and with the overall skill ratings, with correlation coefficients ranging from 0.409 to 0.892 (Table S2).

To examine potential rating bias between raters, we calculated the average z scores for each peer rater. For each video, a z score was calculated as the difference between the score of each rater and the average score of all raters, divided by the standard deviation. None of the raters’ scores were significantly different from the mean.

Participating surgeons were categorized into three groups according to quartiles of skill rating scores: the bottom group (first quartile), middle group (second and third quartiles), and top group (fourth quartile).

Comparisons between groups were made by the ANOVA test on continuous variables Categorical variables were compared by the chi-square test or Fisher’s exact test when appropriate. To account for the cluster effect of patients within surgeons, a multivariable generalized linear mixed models with adjusting the risk factors of patients as fixed effect and the surgeon as random effect were employed for surgical ratings on all complications, MACCE and skill-related complications, respectively.

Odds ratios and corresponding 95% confidence intervals were calculated. The risk factors of patients included age, sex, body-mass index, medical history and left ventricular ejection fraction (LVEF) on baseline. The Kaplan-Meier method was used to determine the cumulative incidence since the surgical procedure of the follow-up outcomes including all-cause mortality, myocardial infarction, stroke and all complications. Differences in survival between groups were evaluated using the log-rank test. A multivariable Cox proportional hazards regression models with adjusting risk factors of patients were employed for surgical ratings on the follow-up outcomes. hazard ratios and corresponding 95% confidence intervals were calculated.

Statistical significance was considered if the two-sided p value was <0.05. All statistical analyses were performed using SPSS statistical software (Version 23, IBM Corp., Armonk, NY).

## RESULTS

### Surgical skill ratings of surgeons and characteristics of patients

Forty-six cardiac surgeons from Fuwai hospital voluntarily participated in this study. All of the surgeon were man. The video of their performance of an isolated CABG was taken by the camera in the operating room. The mean rating of the surgical skill of the 46 surgeons varied substantially, ranging from 3.10 to 4.81 (Figure 1). The 12 surgeons in the bottom quartile had a mean rating of 3.59, as compared to a mean rating of 4.64 for the 11 surgeons in the top quartile. Ratings of each of the seven domains of surgical skill varied to a similar degree (Table 1). Surgeons in the bottom quartile of skill rating, as compared to those in the top quartile, also had longer average operating times for the CABG procedure (183.4±174.0 minutes vs. 165.0±128.5 minutes, p<0.001).

**Figure 1.**
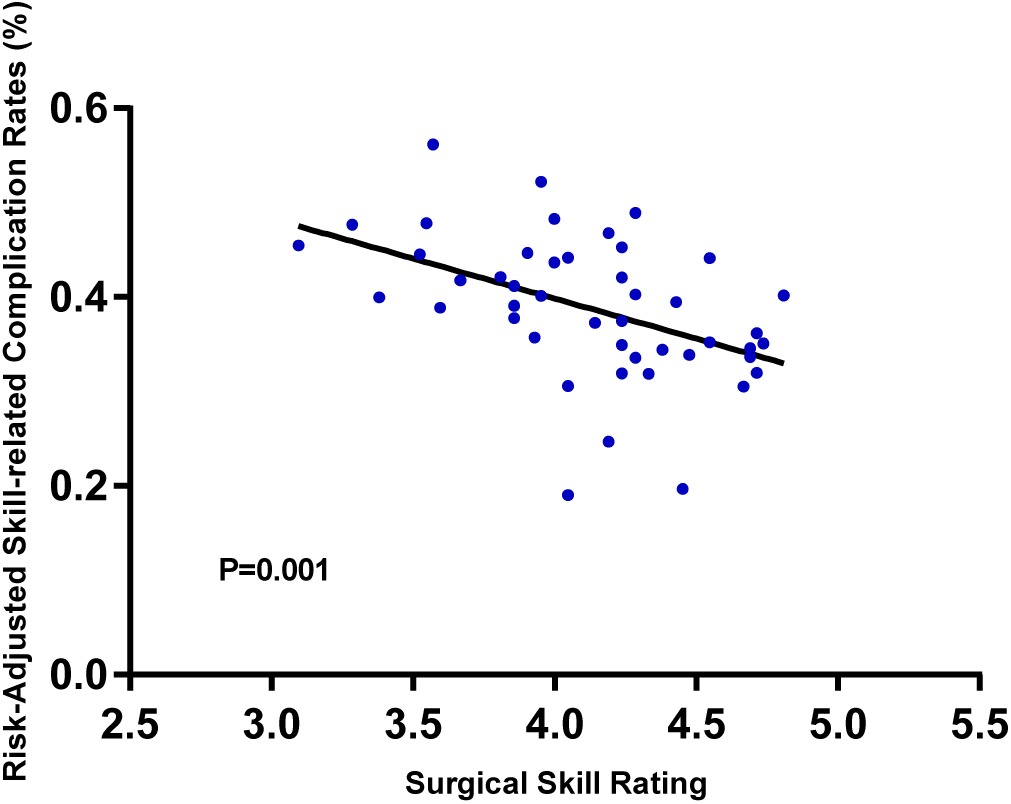
Relationship between Surgical Skill Ratings and Risk-Adjusted Skill-related Complication Rates after Coronary Artery Bypass Graft.

**Table 1.** Characteristics of Surgeons, Patient Volume, and Surgery, According to Peer Ratings of Surgical Skill*.

| Variable | Level of Surgical Skill** |  |  | P Value |
| --- | --- | --- | --- | --- |
|  | Bottom group | Middle group | Top group |  |
| Surgeons (no.) | 11 | 23 | 12 |  |
| Peer ratings of surgical skill |  |  |  |  |
| Gentleness | 3.67±0.35 | 4.15±0.25 | 4.85±0.31 |  |
| Cleanliness | 3.76±0.32 | 4.14±0.26 | 4.61±0.25 |  |
| Motion | 3.19±0.52 | 3.97±0.39 | 4.52±0.40 |  |
| Instrument handling | 3.40±0.45 | 4.20±0.26 | 4.09±0.54 |  |
| Coordinate with nurse | 3.92±0.45 | 4.35±0.38 | 4.65±0.35 |  |
| Coordinate with first assistant | 3.72±0.31 | 4.23±0.29 | 4.58±0.22 |  |
| Fluency and coherence | 3.44±0.41 | 4.04±0.39 | 4.68±0.20 |  |
| Mean | 3.59±0.24 | 4.16±0.15 | 4.64±0.12 |  |
| Age (years) | 49.7±5.7 | 50.9±6.4 | 57.9±4.4 | 0.002 |
| Senior surgeon (n, %) | 3 (25.0) | 13 (56.5) | 10 (90.0) | 0.006 |
| MBTI (n, %) |  |  |  |  |
| Extraversion | 5 (41.7) | 10 (43.5) | 6 (54.5) | 0.790 |
| Sensing | 4 (33.3) | 17 (73.9) | 8 (72.7) | 0.046 |
| Thinking | 10 (83.3) | 22 (95.7) | 6 (54.5) | 0.013 |
| Judging | 4 (33.3) | 6 (26.1) | 3 (27.3) | 0.900 |
| Annual CABG volume (no.) | 41.5±25.0 | 73.7±36.4 | 109.3±57.7 | 0.001 |
| Total CABG volume (no.) | 305.0±359.6 | 705.0±536.4 | 1388.3±791.4 | <0.001 |
| Annual cardiac surgery volume (no.) | 68.9±45.5 | 130.7±62.8 | 229.1±79.8 | <0.001 |
| Total cardiac surgery volume (no.) | 814.7±536.6 | 1625.4±822.7 | 2941.4±1045.6 | <0.001 |
| Patients (no.) | 1355 | 4956 | 3533 |  |
| Mean operating room times (min) | 183.4±174.0 | 171.7±151.3 | 165.0±128.5 | <0.001 |
| ICU stay (hr) | 41.9±52.4 | 41.3±53.1 | 40.6±47.4 | 0.664 |

\*, Each domain of surgical skill was rated on a scale of 1 to 5 (with 1 indicating the skill expected of a cardiac-surgery chief resident and 5 indicating the skill of a master cardiac surgeon). CABG, coronary artery bypass graft. ICU, intensive care unit.
MBTI, Myers-Briggs Type Indicator.
\*\* Quartile 1, 2 and 3 indicates surgeons with the low, medium, and high scores, respectively.

There were no statistically significant differences in patient sex, body-mass index, medical history, or LVEF across quartiles of surgical skill (Table 2). Among patients treated by surgeons in the bottom quartile, compared with those treated by surgeons in the top quartile, there was a younger age (61.1±8.0 vs. 62.2±8.8, p<0.001) and a lower prevalence of smoking (50.5% vs. 55.2%, p=0.011).

**Table 2.** Baseline Characteristics of Patients According to Surgical Skill.

| Variable | Level of Surgical Skill |  |  | P Value |
| --- | --- | --- | --- | --- |
|  | Bottom group<br>(11 surgeons,<br>1355 patients) | Middle group<br>(23 surgeons,<br>4956 patients) | Top group<br>(12 surgeons,<br>3533 patients) |  |
| Mean age (yr) | 61.1±8.0 | 61.7±8.6 | 62.2±8.8 | <0.001 |
| Male sex (n, %) | 1057 (78.0) | 3912 (78.9) | 2771 (78.4) | 0.716 |
| Mean body-mass index* | 25.8±3.2 | 25.8±3.1 | 25.9±3.2 | 0.067 |
| Medical history (n, %) |  |  |  |  |
| Hypertension | 850 (62.7) | 3199 (64.5) | 2329 (65.9) | 0.099 |
| Chronic renal insufficiency | 4 (0.3) | 23 (0.5) | 22 (0.6) | 0.543 |
| Stroke | 110 (8.1) | 480 (9.7) | 334 (9.5) | 0.212 |
| Peripheral Artery Disease | 105 (7.7) | 398 (8.0) | 301 (8.5) | 0.599 |
| Diabetes mellitus | 524 (38.7) | 1953 (39.4) | 1424 (40.3) | 0.523 |
| COPD | 12 (0.9) | 53 (1.1) | 47 (1.3) | 0.344 |
| Congestive heart failure | 7 (0.5) | 23 (0.5) | 20 (0.6) | 0.808 |
| Myocardial infarction | 379 (28.0) | 1519 (30.6) | 1032 (29.2) | 0.107 |
| Atrial fibrillation | 17 (1.3) | 97 (2.0) | 67 (1.9) | 0.222 |
| Cardiac surgery history | 40 (3.0) | 187 (3.8) | 113 (3.2) | 0.199 |
| Smoking | 684 (50.5) | 2642 (53.3) | 1949 (55.2) | 0.011 |
| LVEF (%) | 59.7±7.6 | 59.4±7.5 | 59.2±7.8 | 0.267 |

COPD, chronic obstructive pulmonary disease. LVEF, left ventricular ejection fraction.
\*, The body-mass index is the weight in kilograms divided by the square of the height in meters.

### Association between surgical skill ratings and complication rates

Individual surgeon was an independent predictor for MACCE, skill-related complications and all complications, when adjusted with the patients’ risk factors. Surgical skill ratings were strongly related to overall MACCE, skill-related complications and all complications,. When adjusted with the patients’ risk factors, cases performed by bottom, middle and top quartile rating surgeons had a significant difference on rate of MACCE (9.0%, 6.7% and 6.9%, p=0.016), skill-related complication (43.4%, 38.0% and 34.8%, p=0.005) and all complication rates (52.2%, 48.8% and 47.4%, p=0.002) (Figure 2). After adjustment with the patients’ risk factors, compared to cases performed by the top quartile rating surgeons, those by bottom quartile rating surgeons had 1.59 times the risk in MACCE (95%CI, 1.05-2.40, p=0.029), 1.70 times the risk in skill-related complications (95%CI, 1.33-2.16, p<0.001), and 1.36 times the risk in overall complications (95%CI, 1.09-1.68, p=0.005, Table 3). There was no significant difference on risk in MACCE or overall complications between patients treated by surgeon in the middle group and those in the top group. Compared to cases treated by surgeon in top group, these in the middle group had a 1.26 times the risk in skill-related complications (95%CI, 1.03-1.53, p=0.025, Table 3). Analysis on subgroups of sex and age were performed. The effect of bottom, middle and top quartile rating on MACCE, skill-related complications and overall complications were not affected by sex and age of the patients (Table S3).

**Figure 2.**
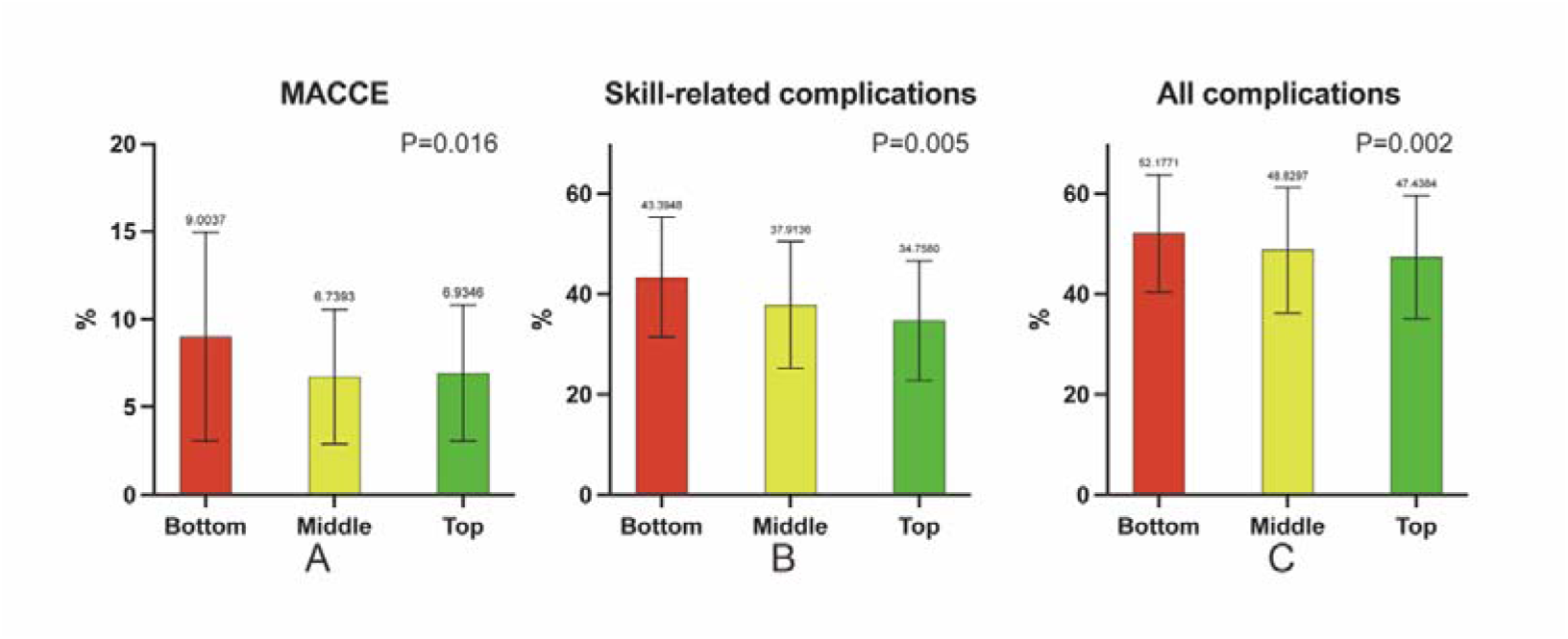
Risk-adjusted Complication Rates According to Skill Ratings.

**Table 3.** Adjusted Risk of Complications According to Surgical Skill.

| Variable, n (%) | OR | 95%CI | P value |
| --- | --- | --- | --- |
| MACCE <sup>a</sup> |  |  |  |
| Top group | 1 |  |  |
| Middle group | 0.96 | 0.68-1.36 | 0.815 |
| Bottom group | 1.59 | 1.05-2.40 | 0.029 |
| Skill-related complications <sup>b</sup> |  |  |  |
| Top group | 1 |  |  |
| Middle group | 1.26 | 1.03-1.53 | 0.025 |
| Bottom group | 1.70 | 1.33-2.16 | <0.001 |
| All complications <sup>c</sup> |  |  |  |
| Top group | 1 |  |  |
| Middle group | 1.12 | 0.94-1.33 | 0.202 |
| Bottom group | 1.36 | 1.09-1.68 | 0.005 |

a, MACCE, major adverse cardiac and cerebrovascular event, included 30-days mortality, myocardial infarction and stroke.

Within an average follow-up of 29.4 months, there were no statistically significant differences in all-cause mortality, myocardial infarction, stroke, chronic kidney injury, revascularization, thrombolysis or all complications above across quartiles of surgical skill ratings (Table 2 and Figure 3). When adjusted with the patients’ risk factors and secondary prevention, surgical skill ratings were not associated with long-terms complications mentioned above (Table S4).

**Figure 3.**
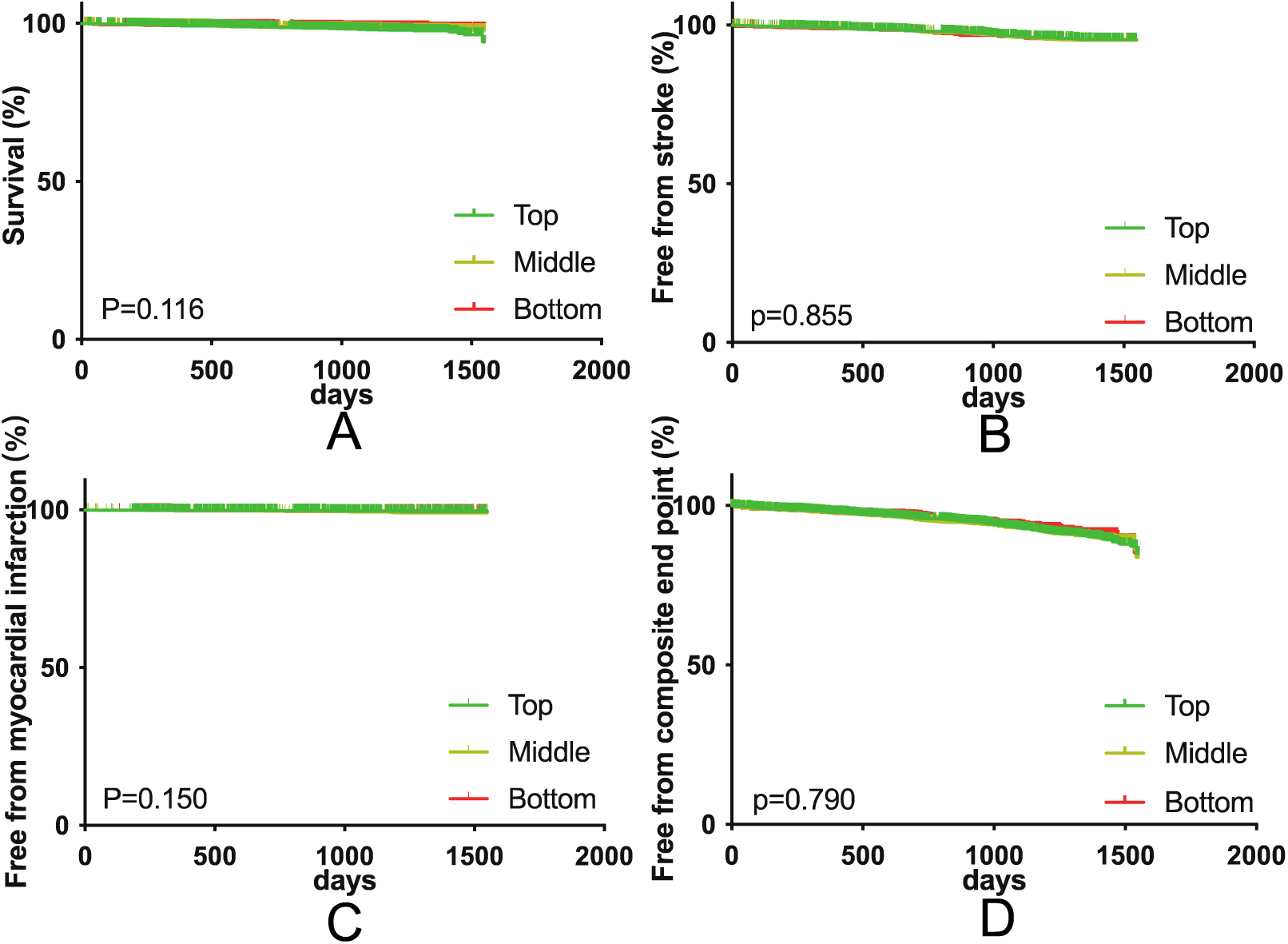
Kaplan-Meier analysis for Long-term Outcomes According to Skill Ratings.

### Relationship between surgical skill ratings and characteristics of surgeons

Surgical skill ratings were strongly related with age, professional titles, MBTI characteristics, and annual procedure volume. Compared with surgeons in the top quartile of skill rating, surgeons in the bottom quartile had a younger age (49.7±5.7 vs. 57.9±4.4, p=0.002), a lower rate of senior professional titles (25% vs. 90%, p=0.006), a lower rate of “sensing” characteristics of MBTI (33.3% vs. 72.7%, p=0.046), a higher rate of “thinking” characteristics of MBTI (83.3% vs. 54.5%, p=0.013), and a lower mean annual volumes of CABG procedures (41.5±25.0 vs. 109.3±57.7, p=0.001).

### Relationship between surgical skill ratings and HRV indices and peer evaluations

During CABG, HRV was analyzed while surgeons performing distal vascular stomas (Figure S1). A significant difference was observed in LF/HF ratio between surgeons in the top, middle and bottom quartile of skill rating (2.3±0.1, 3.0±0.5, and 4.9±1.4 p<0.01). Interestingly, the standard deviation of the LF/HF ratio in surgeons in bottom quartile of skill rating was significantly higher than that in middle and top quartile (1.12±0.67 vs. 0.21±0.15 and 0.17±0.04, p<0.01).

## DISCUSSION

In this large, comprehensive, and contemporary investigation of the relationship between characteristics of senior surgeons and outcomes for CABG, we observed a significant association between the surgical technical skills, as assessed by video-based peer review, and 30-day outcomes. Surgeons with poor surgical skill scores had a 38% risk increase in MACCE and a 29% risk increase in all complications associated with CABGs.

Our findings that variance still exist among senior surgeons highlighted the importance of improvement efforts on surgical skills, which are largely overlooked. Such findings also raise an important question on how to balance the need to gain experience with outcomes. Ideally, guidance from more skilled surgeons should be invited for less skilled surgeons to skill refinement and decision-making. Continuing education may help latter surgeons avoid obtaining skill improvements at the cost of patient safety, as recommended by the Society of Thoracic Surgeons, or through less formal mechanisms.^18–20^

In the current study, the absolute differences in patient characteristics of the three groups (clustering under surgeons with low, medium, and high scores) were small. In general, “high score” cases were sicker and with more comorbidities. It is reasonable because surgeons with high scores can choose patients based on their own judgement and may take on more complex cases. Others have reported that junior surgeons were less likely to perform high-risk procedures and more likely to perform procedures with increasing complexity and rarity as experience accumulated.^21^

Although there was a strong association between surgical skill score and 30-day MACCE/complications, 30-day mortality did not differ throughout the three scoring groups. Possibly, teamwork which has been proven to be effective defense mechanisms, successfully adapted to change for an optimal outcome by avoidanding death and/or near-miss.^19, 20^ However, this factor could not be reflected in this retrospective report. Also, the medium-term outcomes in our study were not different among the three groups even when adjusted by the patients’ risk factors. Medium-term outcomes may be affected by many factors including patients’ characteristcs, surgery techniques and secondary prevention.

This finding echoed the results from previous studies and highlighted the importance of continuous secondary prevention medication.^22–24^

To explore the mechanisms underlying our observation, we used HRV, a widely adopted measurements for mental stress,^25^ to calculate the relative contribution of sympathetic and parasympathetic nervous systems among surgeons during the operation. During distal anastomoses, surgeons scoring high had a relatively lower sympathetic tone. This finding was in line with previous studies demonstrating operation to be a strong stimulus for increased sympathetic tone.^26–28^ Furthermore, our findings showed that the stress level during operations varied among individual surgeons. It seemed that skilled surgeons might be more adaptive to stimulus and thus be more likely to give stable performance during operations.

As surgical skill score was strongly associated with the clinical outcome, improving technical skill should be the key point in training cardiac surgeon. Efforts were made in reforming the training program of cardiac surgeon, focusing on training contents and schedule.^29–31^ This study demonstrated a strong correlation in characteristics of the surgeons and the surgical skill ratings. Training in good qualities of cardiac surgeon should be valued as well as technical training. During the training program, keeping the trainee interested with cardiac surgery, teaching them to operate with ease and deal with crisis calmly, improving their capability of learning, and developing a habit of good preparation and good operation practice should be emphasized.

Our study has several important characteristics:

Firstly, the study was based on a large medical-record abstracted database which complemented existing large studies based on administrative data. ^7–9, 32, 33^ Aside from patients’ information, a comprehensive collection of surgeons’ information including demographics, professional experience, personalities, and HRV during surgery was achieved in the current study.

Secondly, the current study evaluated the surgical skills of experienced cardiac surgeons with a minimum volume of 200. While important, few instruments are available to help evaluate the performance of cardiac surgeons who has passed their training period. The current study provided a feasible and effective way for such assessments.

Thirdly, the study focused on the association between surgical skill and clinical outcomes, which was seldom evaluated.^32, 33^ In the literature, objective evaluation of technical competence has been difficult and often limited to subjective observation and logbooks.^34^ The development of existing objective and valid evaluation tools were largely based on educational research under simulated environment.^10–15^ By linking these tools directly with outcomes in real clinical scenarios, these results yielded from the current study provided evidence for the translation of the emerging evidence into clinical practice.

This study has several limitations.

A single factor of the surgical skill-outcome relationship may not capture all the important processes of care that influence surgical outcomes. In addition to technique aspect, successful outcomes depend on the synergy of a surgeon’s cognitive skills, interpersonal skills, and environmental factors.^35–37^

The composite outcomes were correlated with surgical skill scores. But we found that individual complications were not highly correlated, indicating that different complications may be affected differently with surgical skills. This finding is reasonable and interesting. One would intuitively attribute all the complications to surgical performance. However, in the real world, every staff participating the treatment contributes to the outcomes. Therefore, the quality measures used in the current study might not fully characterize surgical performance and the relationship between surgical skill and individual complication need to be further explored.

In the current study, we employed the methodology of objective structured assessment of technical which requires a task-specific checklist and a global rating scale.^38^ However, it was reported that checklist was not a good discriminator of surgical skills for experienced surgeons, presumably due to the fact that senior surgeons have already been familiar with the steps of procedure and principles of execution.^39^ Therefore, in consideration that the overall experience of the participating surgeons was high and for the convenience of reviewers, we only employed global rating scale in the study.

Finally, we could not adjust our results with the coronary anatomy, coronary functional/physiological indices, and vessels bypassed because this information was not available.

In conclusion, by identifying a significant surgical skill rating-outcome relationship, the study provides a way of systematic, reproducible, and objective skill-based evaluation of senior surgeons in cardiac surgery. This provides an opportunity to improve outcomes by reorganizing case allocation and structure of surgical coverage as well as improving the quality of training programs for cardiac surgeons.

Skill-related complications included post-operative red blood cell infusion, pericardial tamponade, secondary thoracotomy for bleeding, cardiac arrest/ventricular fibrillation, fatal ventricular arrhythmia, myocardial infarction, III° atrial-ventricular block, intra-aortic balloon pump implantation, and aortic dissection. Each diamond in the scatter plot represents 1 of 46 practicing cardiac surgeons.

MACCE, major adverse cardiac and cerebrovascular event. A, MACCE included 30-days mortality, myocardial infarction and stroke. B, Skill-related complications included post-operative red blood cell infusion, pericardial tamponade, secondary thoracotomy for bleeding, cardiac arrest/ventricular fibrillation, fatal ventricular arrhythmia, myocardial infarction, III° atrial-ventricular block, intra-aortic balloon pump implantation, and aortic dissection.C, All complications included 30-days mortality, myocardial infarction, hepatic failure, acute kidney injury, stroke, red blood cell infusion, pericardial tamponade, secondary thoracotomy for bleeding, cardiac arrest/ventricular fibrillation, fatal ventricular arrhythmia, III° atrial-ventricular block, intra-aortic balloon pump implantation, extracorporeal membrane oxygenation implantation, aortic dissection, coma more than 24 hours, dialysis, re-intubation, re-ICU, debridement, mechanical ventilation more than 48h, sepsis, hydrothorax/hydropericardium, pulmonary infection, pulmonary embolism, and atrial fibrillation.

A, free from all-cause mortality. B, free from stroke. C, free from myocardial infarction. D, free from composite endpoint, including all-cause mortality, stroke, myocardial infarction, renal failure, percutaneous transluminal coronary intervention and thrombolysis therapy.

## Data Availability

Data was available.

## Notes

### Competing Interest Statement

The authors have declared no competing interest.

### Author Declarations

The study was approved by the institutional ethics committee of Fuwai Hospital (IRB2012-BG-006)

